# Exploring structural connectomes in children with unilateral cerebral palsy using graph theory

**DOI:** 10.1101/2022.08.12.22278697

**Authors:** Ahmed Radwan, Lisa Decraene, Patrick Dupont, Nicolas Leenaerts, Cristina Simon-Martinez, Katrijn Klingels, Els Ortibus, Hilde Feys, Stefan Sunaert, Jeroen Blommaert, Lisa Mailleux

## Abstract

**Background and objectives:** Brain damage during early development impacts the brain structural network and its coinciding functions. Here, we explored structural brain connectomes in children with unilateral cerebral palsy and its relation to sensory-motor function using a novel semi-automated graph theory analysis, investigating both hemispheres.

**Methods:** In 46 children with spastic unilateral cerebral palsy (mean age 10y7m±2y9m, 27 boys, 25 right-sided hemiplegia; Manual Ability Classification System I=15, II=16, III=15) we assessed upper limb somatosensory (two-point discrimination and stereognosis) and motor function (grip force, Assisting Hand Assessment and Jebsen-Taylor Hand Function Test). We collected multi-shell diffusion-weighted, T1-weighted and T2-FLAIR MRI and performed transcranial magnetic stimulation to identify the corticospinal tract (CST) wiring pattern. Structural connectomes were constructed using Desikan-Killiany parcellations with Virtual Brain Grafting and FreeSurfer, and Multi-Shell Multi-Tissue Constrained Spherical Deconvolution diffusion modelling with anatomically constrained tractography. Graph metrics (characteristic path length, global/local efficiency and clustering coefficient) were calculated for the whole brain, the ipsilesional/contralesional hemisphere, and the full/ipsilesional/contralesional sensory-motor network, and were compared between lesion types (periventricular white matter (PWM)=28, cortical and deep grey matter (CDGM)=18) and CST-wiring patterns (ipsilateral=14, bilateral=14, contralateral=12, unknown=6) using ANCOVA with age as covariate. We used elastic-net regularized regression to investigate how graph metrics, lesion volume, lesion type, CST-wiring pattern and age predicted sensory-motor function.

**Results:** In both the whole brain and subnetworks, we observed a hyperconnectivity pattern in children with CDGM-lesions compared to PWM-lesions, with higher clustering coefficient (p=[<0.001-0.047], 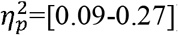), characteristic path length (p=0.003, 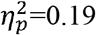) and local efficiency (p=[0.001-0.02], 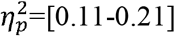), as well as a faster decrease in global efficiency with age (p=[0.01-0.04], 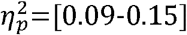). No differences were found between CST-wiring groups. In general, good predictions of sensory-motor function were obtained with elastic-net regression (R^2^=0.40-0.87). For motor function, the CST-wiring pattern was identified as the strongest predictor. For somatosensory function all independent variables contributed equally to the model.

**Discussion:** With this study, we demonstrated the feasibility and potential of structural connectomes using graph theory analysis in understanding disease severity and brain development in children with unilateral cerebral palsy. Hence, this exploratory study could support and direct future research.

## Introduction

The complexity of the human brain is thought-provoking and still not fully understood. The human brain involves multiple, closely intertwined, networks that allow us to perform highly skilled movements and cognitive processes. Advancements in Diffusion magnetic resonance imaging (dMRI) and tractography techniques have enabled the investigation of both micro- and macro-structural changes in the brain’s white matter, enriching our understanding of the complex brain architecture and its development.

A brain lesion occurring early in life can disrupt the typical development of these brain networks and even result in alteration of the brain’s anatomical architecture, as is the case for children having cerebral palsy (CP). CP is the most common cause of disability in children^1^, and is defined as “*a group of permanent disorders of the development of movement and posture, causing activity limitation, that are attributed to non-progressive disturbances that occurred in the developing fetal or infant brain; the motor disorders of CP are often accompanied by disturbances of sensation, perception, cognition, communication, and behaviour, by epilepsy, and by secondary musculoskeletal problems*”.^2^ In one third of these children, sensory-motor impairments are predominantly present on one side of the body, also referred to as unilateral CP (uCP).^3^

Multiple studies have used Diffusion MR imaging in an attempt to improve our understanding of structure-function relationships in children with uCP, in particular regarding upper limb functionality, indicating a relation between the severity of the underlying brain damage and upper limb sensory-motor function.^4,5^ However, most studies focused on specific regions of interest, mostly part of the primary sensory-motor areas, while motor actions go beyond activity in the primary sensory-motor network and involve multiple networks across the brain.^5^ Moreover, compared to typically developing children, studies in CP using whole brain dMRI structural connectivity analyses revealed a reduced white matter connectivity not only in sensory-motor regions, but also in non-motor areas.^6,7^ This underlines the need for further study of the brain structural networks which would allow us to investigate how the structural network reorganises following brain damage early in life and how this relates to sensory-motor function.

Indeed, we could expect that in the case of CP, brain changes occur on a network scale through individual neuroplastic processes. Graph theory (GT) analysis can investigate the brain network characteristics using dMRI tractography, providing information on both global and local integration in the brain.^8,9^ Also, it can reveal changes in brain organisation that extend beyond the injured brain areas. Such information is essential for understanding global and long-term functional outcomes of focal structural brain injury. Moreover, contrary to other analysis techniques, GT-analyses of structural connectomes are not dependent on a single region or structure that may be distorted or absent due to severe brain pathology in these children. However, the construction of structural connectome does require an accurate parcellation map from which to derive the nodes of the network. Yet, brain parcellation is challenging in populations with large lesions and notable distortion of the normal brain structure, as often encountered in children with CP. Therefore, we applied a lesion inpainting method^10^ to minimise parcellation errors, which allowed us to investigate the lesioned hemisphere and include patients with large lesions who would have otherwise been excluded, without interpreter bias. Additionally, we used a tailored processing pipeline that accounted for the presence of lesions in all analysis stages.

In this study, we will explore the structural connectomes of the dominant (contralesional) and non-dominant (ipsilesional) hemisphere in children with uCP. using graph theory analysis. We will first investigate whether structural brain connectomes differ across uCP-specific classification groups (i.e. lesion type and corticospinal tract (CST) wiring pattern). Regarding lesion type, we hypothesised a more damaged structural brain network in children with late-onset lesions predominantly affecting grey matter structures (i.e. cortical and deep grey matter, CDGM) compared to children with early-onset lesions predominantly affecting the white matter (i.e. periventricular white matter, PWM). Regarding the CST-wiring pattern, we hypothesised that children in whom ipsilateral CST-projections control the impaired hand (i.e. ipsilateral and bilateral CST-wiring pattern), would have a more damaged structural brain network, compared to children with a contralateral CST-wiring pattern. Secondly, we explored the added value of GT-measures in predicting upper limb sensory-motor function along with other neurological factors (i.e. lesion type, CST-wiring pattern and lesion volume).

## Methods

### Participants

Children with a predominantly spastic type of uCP, aged 5 to 15 years were recruited between May 2014 and April 2017 via the CP-care program of the University Hospitals Leuven (Belgium). Exclusion criteria were: (1) botulinum toxin-A injections 6 months prior to testing, (2) upper limb surgery in the past 2 years and (3) any contraindications for MRI.

All children underwent an upper limb evaluation, including a clinical assessment of motor and sensory impairments, and an evaluation of bimanual performance and unimanual capacity at the Clinical Motion Analysis Laboratory of the University Hospitals Leuven. Children were assessed by three well-trained physiotherapists who were routinely involved in the clinical evaluation of children with CP. On the same day or within a time interval of maximum four months, all children underwent a scanning protocol including structural and dMRI scans, as well as transcranial magnetic stimulation (TMS) to determine the CST-wiring pattern.

The study was approved by the Ethical Committee of the University Hospitals Leuven (S55555, S56513) and parental written informed consent was obtained for all children prior to participation, according to the Declaration of Helsinki. Children aged 12 years or above were additionally asked for their written assent prior to participation.

### Clinical assessment

The evaluation of sensory-motor impairments included grip force and somatosensory function (i.e. two-point discrimination and stereognosis). Grip force was evaluated with the Jamar dynamometer (Lafayette Instrument Company, Lafayette, IN, USA), using the mean of three maximum contractions of each hand. Two-point discrimination and stereognosis were assessed according to Klingels et al.^11^ Briefly, two-point discrimination was examined distally at the index finger using an aesthesiometer to identify the minimal distance at which one or two points could be correctly distinguished. Stereognosis was evaluated via tactile identification of six familiar objects.

At activity level, bimanual performance was assessed using the Assisting Hand Assessment (AHA).^12,13^ During a video-recorded semi-structured play session, the AHA evaluates the spontaneous use of the impaired hand during bimanual activities. Afterwards, 22 items were scored and converted to 0–100 logit-based AHA units. Unimanual capacity was assessed at both hands with the Jebsen-Taylor Hand Function Test (JTHFT), evaluating movement duration during the execution of six unimanual tasks.^14^

### MRI acquisition

All MR images were acquired using the same scanner (3T Philips Achieva, 32-channel phased-array head coil) and scanning protocol. To limit motion, a familiarisation protocol was used with children up to 10 years of age,^15^ and all children were able to watch a movie during scanning. Multi-shell diffusion-weighted images were acquired with spatial resolution = 2.5×2.5×2.5mm^3^, TR/TE = 7800/90ms, Flip angle = 90° phase encoding = AP, b-values = 0/700/1000/2800 with respectively 10/25/40/75 uniformly distributed gradient directions, in-plane parallel acceleration factor (SENSE)^16^ = 2.5, acquisition matrix = 96 × 96 × 50. High-resolution T1-weighted images (MPRAGE) were acquired with spatial resolution = 1.2x.98x.98mm^3^, TR/TE = 9.6/4.6 ms, Flip angle = 8°, acquisition matrix = 160 × 256 × 256, and 3D turbo spin-echo. T2-weighted fluid-attenuated inversion recovery images (T2-FLAIR) were acquired with spatial resolution = .71x.71×1.2mm^3^, TE/TR/TI = 415/4800/1650ms, Flip angle = 90°, acquisition matrix 352 × 352 × 299.

### Image processing

All images were semi-automatically processed by combining publicly available toolboxes and in-house developed scripts (using Bash and Matlab v2020b). Below we briefly discuss the processing steps and give a visual overview of the processing pipeline (Figure 1). A detailed description is given in the supplementary methods.

**Figure 1.**
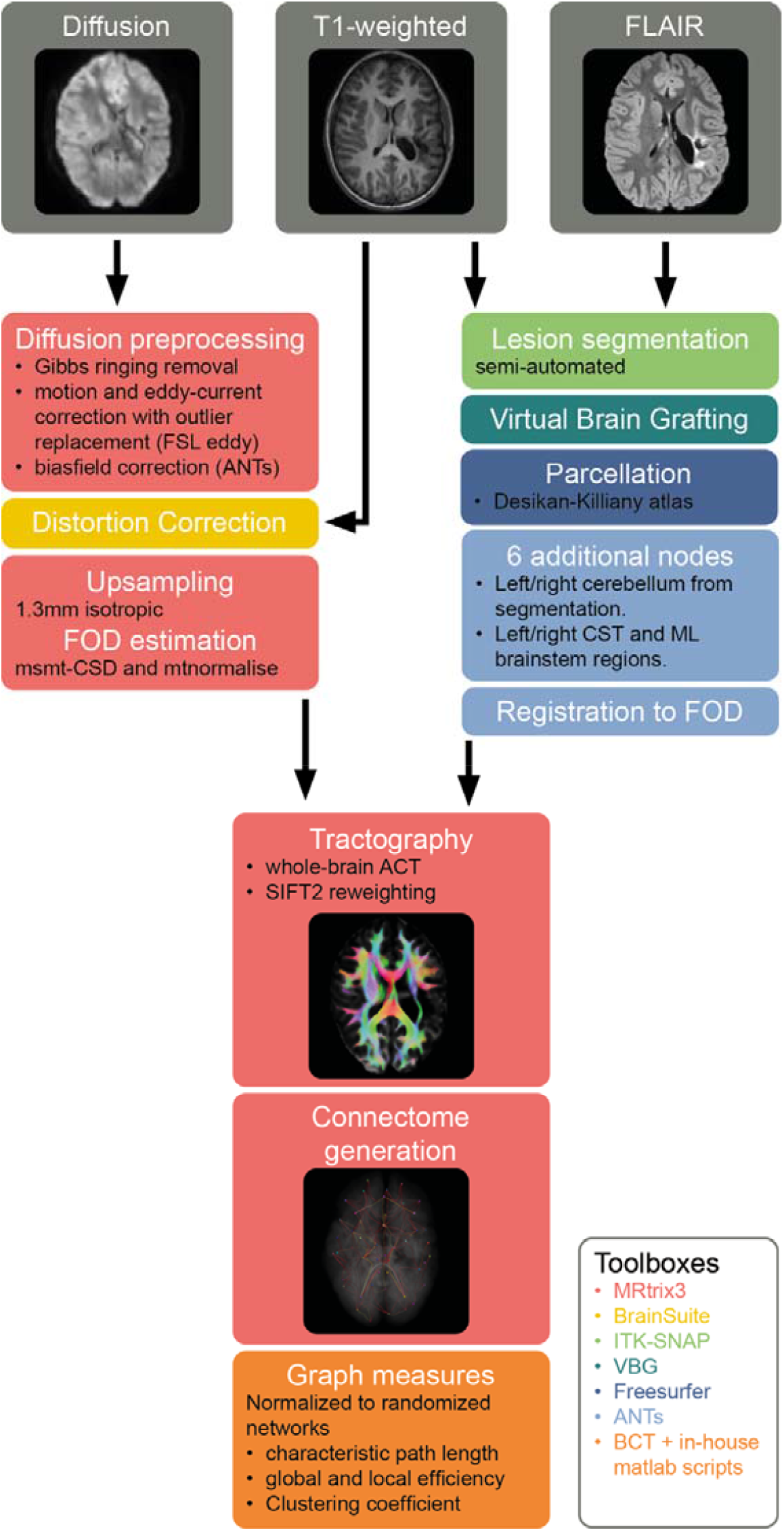
Overview of the imaging analysis pipeline, color-coded by software packages used. ACT: anatomically constrained tractography. BCT: brain connectivity toolbox. CST: corticospinal tract. FOD: fibre orientation distribution. ML: Medial lemniscus. msmt-CSD: multi-shell multi-tissue constrained spherical deconvolution. SIFT: spherical-deconvolution informed filtering of tractograms. VBG: virtual brain grafting.

Manual lesion segmentation was done in ITK-snap^17^ (v3.8.0) by a neuroradiologist (AR) using the T1 and T2-FLAIR weighted images, and lesion volumes were calculated. Virtual Brain Grafting^10^ (VBG v0.31) was used to generate synthetic lesion-free images from the T1-weighted images and lesion masks, which were used for whole brain structural parcellation with FreeSurfer^18^ (v6.0) recon-all. The resulting Desikan-Killiany^19^ parcellation maps were transformed to the mean fibre orientation distribution (FOD) image using ANTs^20,21^ (v2.3.1) affine registration followed by nonlinear registration with a mutual information cost-function, and appended with four brainstem regions from the UKBB volumetric white matter bundles^22^ atlas for the bilateral corticospinal tracts (CST) and medial lemnisci (ML), henceforth referred to as the modified Desikan-Killiany parcellation maps.

Diffusion-weighted images were preprocessed using an MRtrix3 (v3.0) based^23^ pipeline including, image denoising^24^, gibbs ringing correction^25^, motion and eddy current correction with FSL (v6.0.1) Eddy^26^, and ANTs^20,21^ N4 bias-field correction^27^. The T1-weighted images were bias-field corrected and used for EPI distortion correction in the BrainSuite Diffusion Pipeline (Bhushan et al., 2015, 2012) (BDP v19a).^27,28^ Subject-level tissue specific constrained spherical deconvolution (CSD) response functions were generated using multi-shell multi-tissue CSD and a group-level average tissue-specific response function was calculated in MRTrix3.^29^ Individual FOD maps were generated based on the group-average response functions, and normalised for multiple-tissues. Whole-brain tractograms were generated for each subject (iFOD2, 10 million fibres, minimum/maximum length = 5/300mm, maximum angle = 45°), using anatomically constrained tractography^30^, including amygdalae and hippocampi as subcortical grey matter and SIFT2 re-weighting of streamlines.^30,31^

Structural connectomes were constructed with MRTrix3 for each subject with TCK2connectome^32^ using the whole brain, SIFT2-weighted, tractograms and modified Desikan-Killiany parcellations^19^. GT-analysis was performed using in-house developed MATLAB scripts and the Brain Connectivity toolbox (v2019-03-03). GT-measures of characteristic path length, global and local efficiency and clustering coefficient were calculated on multiple levels, namely: the whole brain (using all 88 nodes), separate ipsilesional (using all 41 supratentorial ipsilesional nodes) and contralesional (using all 41 supratentorial contralesional nodes) hemispheres, as well as the full sensory-motor network (SMN, 22 nodes, see Supplementary Material Table A), and ipsilesional and contralesional SMN (each 11 nodes, see Supplementary Material Table A). Self-connections were removed from all connectomes. Nodes where less than 1000 streamlines arrived were defined as disconnected nodes and were removed from the connectome. Subsequent, disconnected nodes in a particular subnetwork, having no connections to other nodes within the subnetwork, were removed from that subnetwork. All edge weights were divided by the maximum edge weight. Characteristic path length and global efficiency were calculated using Dijkstra’s algorithm^33^, with the connection-length matrix defined by the inverse edge weights. Clustering coefficient and local efficiency measures were calculated as recommended by Wang et al.^34^ Briefly, characteristic path length measures the average distance between any two nodes in the network. Clustering coefficient expresses the tendency of a network to be organised in densely connected groups (clusters) and is characterized by the connectivity between neighbouring nodes. Global efficiency refers to the mean inverse distance between two nodes in the network. Similarly, local efficiency is a measure of the mean inverse distance between the neighbours of a node, excluding the node itself. For each subnetwork, 100 random graphs were calculated by randomly permuting the edges, while keeping the connectome symmetry, the zero-weight of the self-connections and excluding graphs with disconnected nodes. All graph measures were normalised by dividing the original graph measure by the median graph measure of the equivalent random networks.

### Transcranial Magnetic Stimulation

We performed single-pulse TMS to identify the underlying CST-wiring pattern using a MagStim 200 Stimulator (Magstim Ltd., Whitland, Wales, UK) with a 70 mm figure-eight coil and a Bagnoli electromyography system (Delsys Inc., Natick, MA, USA). After identifying the hotspot and the resting motor thresholds, motor evoked potentials were elicited and recorded for both adductor pollicis brevis muscles to identify the CST-wiring pattern (contralateral, bilateral or ipsilateral). A more detailed description can be found elsewhere.^35^

### Statistical analysis

Descriptive statistics were collected including age, sex and side of uCP. One-way analyses of covariance (ANCOVA) were used to investigate the difference of the GT-measures between lesion types (PWM and CDGM) and between CST-wiring groups (contralateral, bilateral, ipsilateral) with age as a covariate. If a non-significant interaction was found between age and the clinical group, only the model with the main effects was retained. Normal distribution of the residuals was reviewed and confirmed for each fitted model.^36^ Partial eta squared 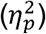 values were calculated to indicate effect sizes and interpreted as small (0.01–0.06), medium (0.06–0.14), and large (>0.14).^37^ For the CST-wiring pattern, a Bonferroni post hoc test, for comparing three CST-groups, was implemented with a corrected p-value (α=0.05). SPSS Statistics version 27.0 (IBM, Armonk, New York, USA) was used.

Next, we explored the value of adding GT-measures to other neurological factors (lesion volume, lesion type and CST-wiring pattern) in predicting upper limb sensorimotor function. This was done through elastic-net regularized regression which optimizes between ridge regression (L2) which shrinks coefficients, and LASSO (L1) regression which excludes predictors that do not add to the model.^38^ The amount of ridge or LASSO regression is expressed by a variable alpha which ranges from 0 (only ridge regression) to 1 (only LASSO). The models were evaluated using R-squared (R^2^) and root-mean-square error (RMSE). After taking the average of 100 cross-validation errors for each alpha and lambda combination, the highest alpha and lambda with a mean cross-validation error that fell within one standard error of the combination with the lowest mean cross-validation error was used. This resulted in the selection of the best, most parsimonious model. The continuous variables (GT measures, lesion volume and age) were standardized so that the estimates can be interpreted as effect sizes. Further, dummy variables were created for the categorical variables (lesion type and CST-wiring pattern). The R^2^ was interpreted according to Cohen^39^ as weak (0.02), moderate (0.13) or substantial (0.26). The effect sizes of the individual predictors were interpreted according to Cohen’s |d| with values <0.1 as tiny, values between 0.1 and 0.2 as very small, between 0.2 and 0.5 as small, between 0.5 and 0.8 as moderate, between 0.8 and 1.2 as large, between 1.2 and 2.0 as very large and >2.0 as huge.^40^ This analysis was performed with R (version 4.1.1).

### Data availability statement

The anonymized data that support the findings of this study, as well as related documents, are available from the corresponding author upon reasonable request.

## Results

### Participants

Fifty-five children with spastic uCP were included. Average age at time of the MRI assessment was 10 years and 7 months (SD 2 years and 9 months; age range 5 years 6 months to 15 years and 10 months). Regarding lesion type, one child was classified as having a cortical maldevelopment, six children had an acquired brain lesion and two children presented with a normal structural MRI scan. These children were excluded from further analyses as these small groups do not allow statistical comparison. Of the remaining 46 children, 28 were classified in the PWM group and 18 in the CDGM group. Regarding the type of the CST-wiring pattern, 12 children had a contralateral CST-wiring pattern (10 PWM and 2 CDGM), 14 had bilateral CST-projections (7 PWM, 7 CDGM) and 14 ipsilateral CST-projections (8 PWM, 6 CDGM). Due to the presence of epilepsy (N=3) or due to refusal (N=3), we were unable to perform the TMS assessment in these children. Table 1 displays an overview of the participant’s characteristics. In addition, a detailed overview of the descriptive and clinical characteristics according to lesion types and CST-wiring pattern is provided in Supplementary Material (Table B and C).

**Table 1.**
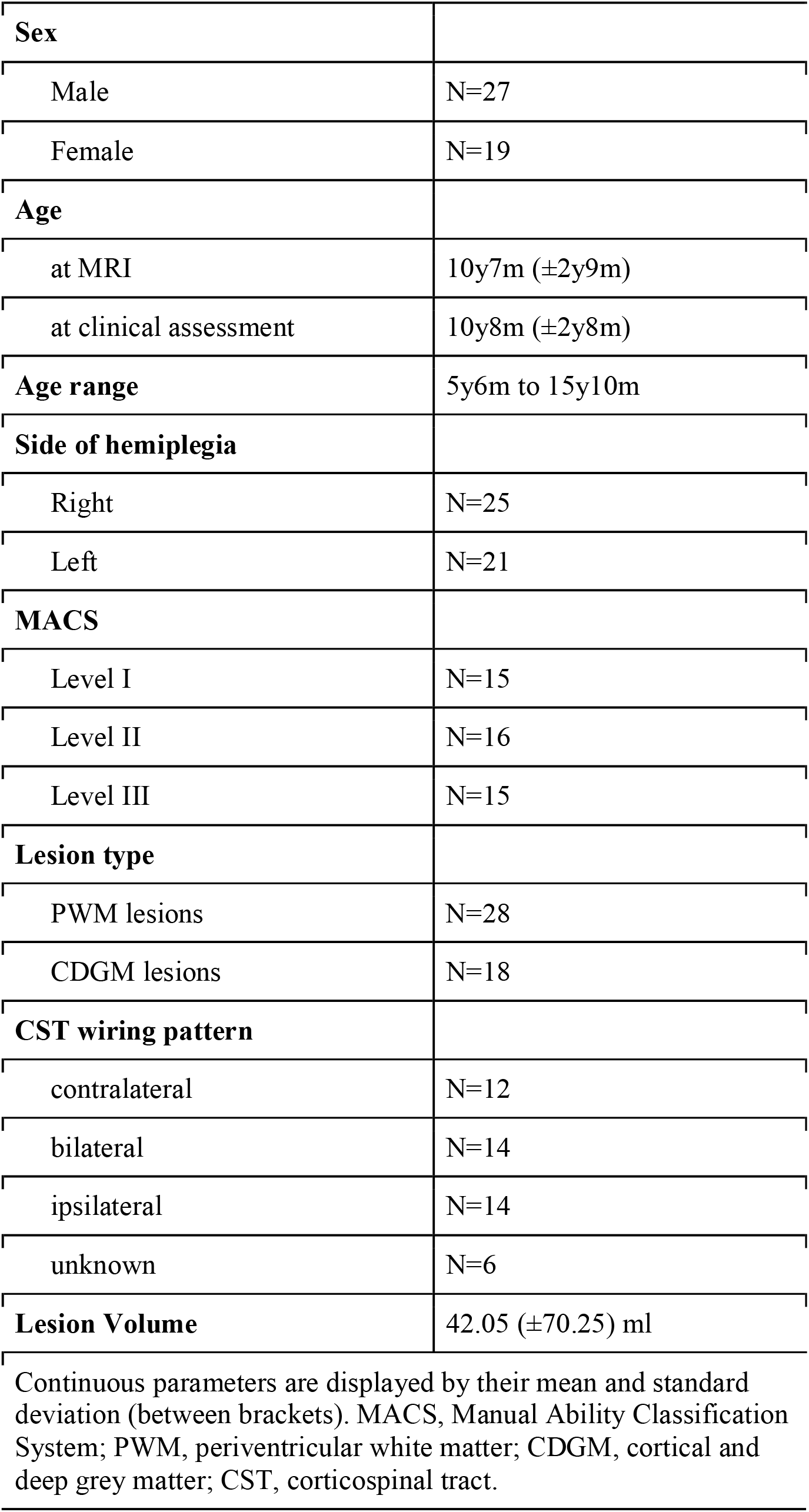
Participant’s characteristics.

|  |  |
| --- | --- |
| **Sex** |  |
| Male | N=27 |
| Female | N=19 |
| **Age** |  |
| at MRI | 10y7m ($\pm 2y9m$) |
| at clinical assessment | 10y8m ($\pm 2y8m$) |
| **Age range** | 5y6m to 15y10m |
| **Side of hemiplegia** |  |
| Right | N=25 |
| Left | N=21 |
| **MACS** |  |
| Level I | N=15 |
| Level II | N=16 |
| Level III | N=15 |
| **Lesion type** |  |
| PWM lesions | N=28 |
| CDGM lesions | N=18 |
| **CST wiring pattern** |  |
| contralateral | N=12 |
| bilateral | N=14 |
| ipsilateral | N=14 |
| unknown | N=6 |
| **Lesion Volume** | 42.05 ($\pm 70.25$) ml |

Continuous parameters are displayed by their mean and standard deviation (between brackets). MACS, Manual Ability Classification System; PWM, periventricular white matter; CDGM, cortical and deep grey matter; CST, corticospinal tract.

### Graph theory measures across lesion type and CST-wiring pattern groups

Figure 2 shows the resulting connectomes for a child with a PWM lesion (panel a) and a child with an extensive CDGM lesion (panel b). As can be visually depicted, the large extent of the lesion in panel b led to multiple nodes in the connectomes to be disconnected, which appeared in 12/46 participants (median=2 disconnected nodes, range=[1-13]) in this study. By removing these disconnected nodes from the connectome, these participants could be successfully included in all analyses.

**Figure 2.**
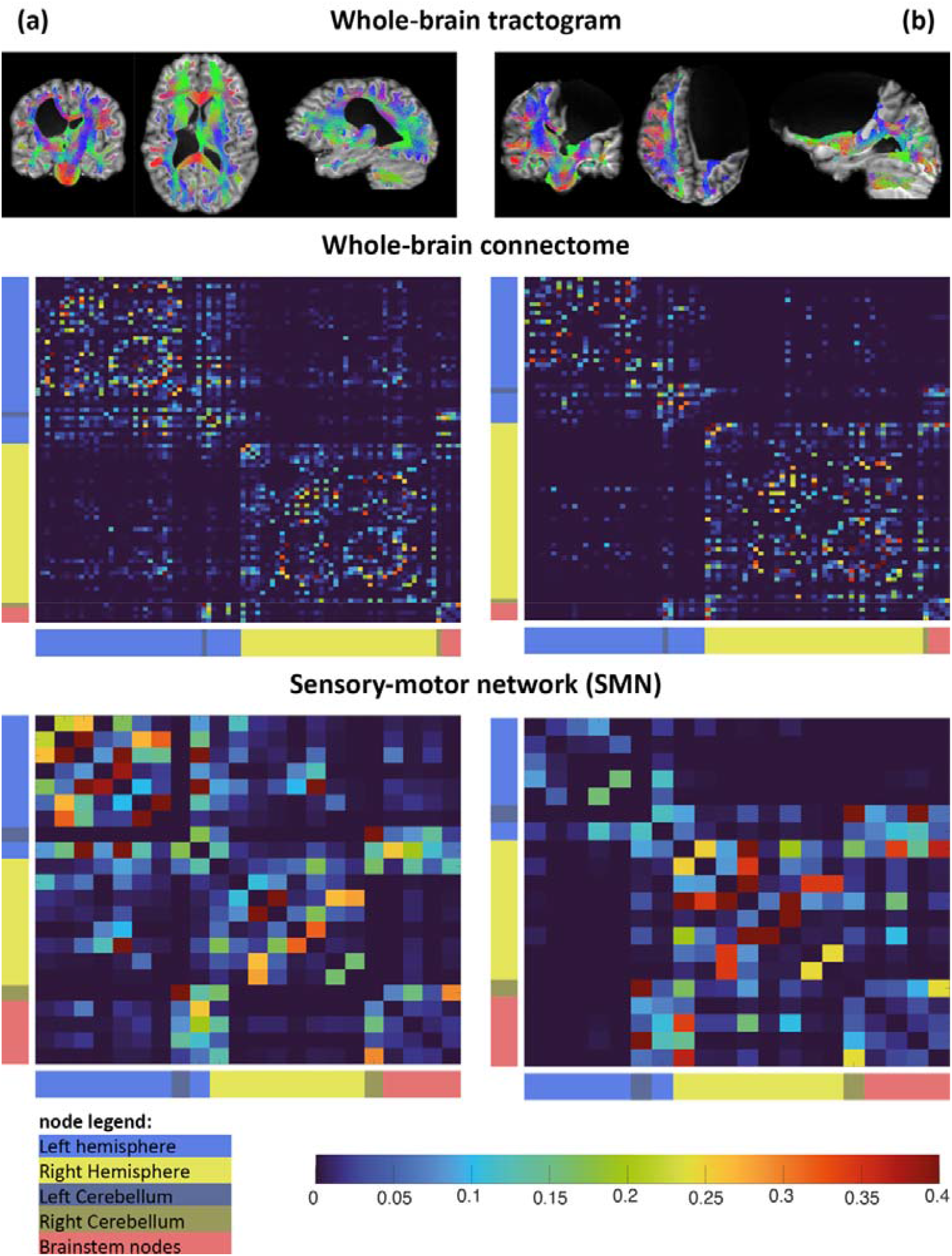
Tractograms, whole-brain connectomes and SMN networks of two participants. Figure 2a shows the connectome of a child with a right lateralized white matter lesion. Figure 2b shows the connectome of a child with an extensive left lateralized cortical and deep grey matter lesion. All edge weight values are normalized to the maximum edge weight in the connectome.

For *lesion type*, significant main effects were found with higher values in the CDGM group compared to the PWM group with effect sizes ranging from medium to large (Table 2) for following parameters. Clustering coefficient was significantly higher in the whole brain (p=0.048; 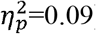); ipsilesional hemisphere (p<0.001, 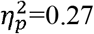), full SMN (p=0.001, 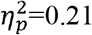), and ipsilesional SMN (p=0.01, 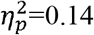). A higher characteristic path length was found in the full SMN (p=0.003, 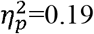). Local efficiency was significantly higher in the whole brain (p=0.01, 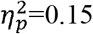), full SMN (p=0.001, 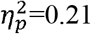) and contralesional SMN (p=0.02, 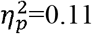). Additionally, a significant difference was found for global efficiency in the whole brain (p=0.01, 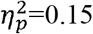), contralesional hemisphere (p=0.03, 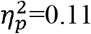) and contralesional SMN (p=0.04, 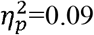), but with an interaction effect of age, indicating that global efficiency decreases more with age in children with CDGM lesions compared to children with PWM lesions. Another interaction effect with age was found for characteristic path length in the whole brain (p=0.01, 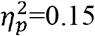), which increased more with age in children with CDGM lesions. The interaction effects are visualised in Supplementary Figures S1-S5.

**Table 2.** Normalized Graph theory measures across lesion types.

| Table 2. Normalized Graph theory measures across lesion types. |  |  |  |  |  |  |  |  |
| --- | --- | --- | --- | --- | --- | --- | --- | --- |
| Graph measures | X(SD) |  | Interaction |  | Lesion Type |  | Age |  |
| | PWM | CDGM | F | P ($\eta_p^2$) | F | P ($\eta_p^2$) | F | P ($\eta_p^2$) |
| **Whole brain** |  |  |  |  |  |  |  |  |
| Char. PL. | 1.34<br>(0.09) | 1.50<br>(0.24) | 7.25 | **0.01**<br>**(0.15)** | 2.69 | 0.11<br>(0.06) | 27.49 | **<0.001**<br>**(0.40)** |
| Cluster Coeff. | 2.53<br>(0.30) | 2.73<br>(0.32) | 0.96 | 0.33<br>(0.022) | 4.16 | **0.048**<br>**(0.09)** | 0.34 | 0.56<br>(0.008) |
| Global eff. | 0.87<br>(0.02) | 0.86<br>(0.04) | 7.26 | **0.01**<br>**(0.15)** | 4.92 | **0.03**<br>**(0.11)** | 16.86 | **<0.001**<br>**(0.29)** |
| Local eff. | 8.46<br>(2.22) | 10.3<br>(2.26) | 0.07 | 0.79<br>(0.002) | 7.27 | **0.01**<br>**(0.15)** | 3.26 | 0.08<br>(0.07) |
| **Ipsilesional hemisphere** |  |  |  |  |  |  |  |  |
| Char. PL. | 1.34<br>(0.11) | 1.35<br>(0.19) | 3.78 | 0.06<br>(0.08) | 0.001 | 0.97<br>(0.00) | 7.03 | **0.01**<br>**(0.14)** |
| Cluster Coeff. | 1.99<br>(0.12) | 2.25<br>(0.31) | 3.23 | 0.08<br>(0.07) | 15.53 | **<0.001**<br>**(0.27)** | 0.73 | 0.40<br>(0.02) |
| Global eff. | 0.89<br>(0.02) | 0.89<br>(0.04) | 2.00 | 0.16<br>(0.05) | 0.006 | 0.94<br>(0.00) | 0.00 | 0.98<br>(0.00) |
| Local eff. | 4.39<br>(0.95) | 5.84<br>(3.74) | 0.00 | 0.99<br>(0.00) | 3.45 | 0.07<br>(0.07) | 2.26 | 0.14<br>(0.05) |
| **Contralesional hemisphere** |  |  |  |  |  |  |  |  |
| Char. PL. | 1.37<br>(0.17) | 1.43<br>(0.28) | 1.79 | 0.19<br>(0.04) | 0.42 | 0.52<br>(0.01) | 18.36 | **<0.001**<br>**(0.20)** |
| Cluster Coeff. | 1.95<br>(0.13) | 1.94<br>(0.16) | 1.55 | 0.22<br>(0.04) | 0.14 | 0.71<br>(0.003) | 0.42 | 0.52<br>(0.01) |
| Global eff. | 0.88<br>(0.03) | 0.90<br>(0.03) | 5.28 | **0.03**<br>**(0.11)** | 7.00 | **0.01**<br>**(0.14)** | 11.47 | **0.002**<br>**(0.21)** |
| Local eff. | 3.97<br>(0.78) | 4.25<br>(0.63) | 0.00 | 0.99<br>(0.000) | 1.31 | 0.26<br>(0.03) | 5.40 | **0.02**<br>**(0.11)** |
| **Full SMN** |  |  |  |  |  |  |  |  |
| Char. PL. | 1.15<br>(0.14) | 1.65<br>(0.85) | 0.22 | 0.65<br>(0.005) | 9.96 | **0.003**<br>(0.19) | 0.50 | 0.48<br>(0.01) |
| Cluster Coeff. | 1.65<br>(0.20) | 1.89<br>(0.27) | 0.60 | 0.44<br>(0.01) | 11.55 | **0.001**<br>(0.21) | 0.45 | 0.50<br>(0.01) |
| Global eff. | 0.94<br>(0.05) | 0.93<br>(0.07) | 0.91 | 0.35<br>(0.02) | 0.08 | 0.78<br>(0.002) | 0.25 | 0.62<br>(0.006) |
| Local eff. | 3.28<br>(0.71) | 4.15<br>(1.11) | 0.04 | 0.85<br>(0.001) | 11.66 | **0.001**<br>**(0.21)** | 2.79 | 0.10<br>(0.06) |
| **Ipsilesional SMN** |  |  |  |  |  |  |  |  |
| Char. PL. | 1.13<br>(0.14) | 1.38<br>(0.65) | 0.33 | 0.57<br>(0.008) | 3.59 | 0.06<br>(0.08) | 0.07 | 0.79<br>(0.002) |
| Cluster Coeff. | 1.46<br>(0.09) | 1.61<br>(0.28) | 1.75 | 0.19<br>(0.04) | 6.97 | **0.01**<br>**(0.14)** | 0.03 | 0.87<br>(0.00) |
| Global eff. | 0.95<br>(0.05) | 0.96<br>(0.08) | 2.46 | 0.12<br>(0.06) | 0.20 | 0.66<br>(0.01) | 0.65 | 0.65<br>(0.01) |
| Local eff. | 2.73<br>(1.93) | 3.68<br>(4.73) | 0.17 | 0.68<br>(0.004) | 0.96 | 0.33<br>(0.02) | 0.19 | 0.66<br>(0.00) |
| **Contralesional SMN** |  |  |  |  |  |  |  |  |
| Char. PL. | 1.16<br>(0.26) | 1.08<br>(0.13) | 1.14 | 0.29<br>(0.03) | 1.58 | 0.22<br>(0.04) | 0.35 | 0.56<br>(0.00) |
| Cluster Coeff. | 1.47<br>(0.08) | 1.44<br>(0.08) | 0.42 | 0.83<br>(0.001) | 1.95 | 0.17<br>(0.04) | 2.86 | 0.10<br>(0.06) |
| **Global eff.** | 0.96<br>(0.04) | 0.98<br>(0.05) | 4.32 | **0.04**<br>**(0.09)** | 5.78 | **0.02**<br>**(0.12)** | 0.07 | 0.79<br>(0.002) |
| **Local eff.** | 2.10<br>(0.49) | 2.68<br>(1.24) | 0.86 | 0.36<br>(0.02) | 5.48 | **0.02**<br>**(0.11)** | 1.75 | 0.19<br>(0.04) |

Char. PL., characteristic path length; coeff, coefficient; eff, efficiency; SMN, sensory-motor network; PWM, periventricular white matter; CDGM, cortical and deep grey matter.

No significant differences were found across *CST-wiring pattern* groups (p>0.08, 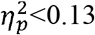), except for characteristic path length in the contralesional hemisphere (p=0.006, 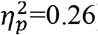) and in the ipsilesional SMN (p=0.02, 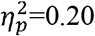), but with an interaction effect of age, and without significant post-hoc differences (see Table 3 and Supplementary Figures S6-S7).

**Table 3.**
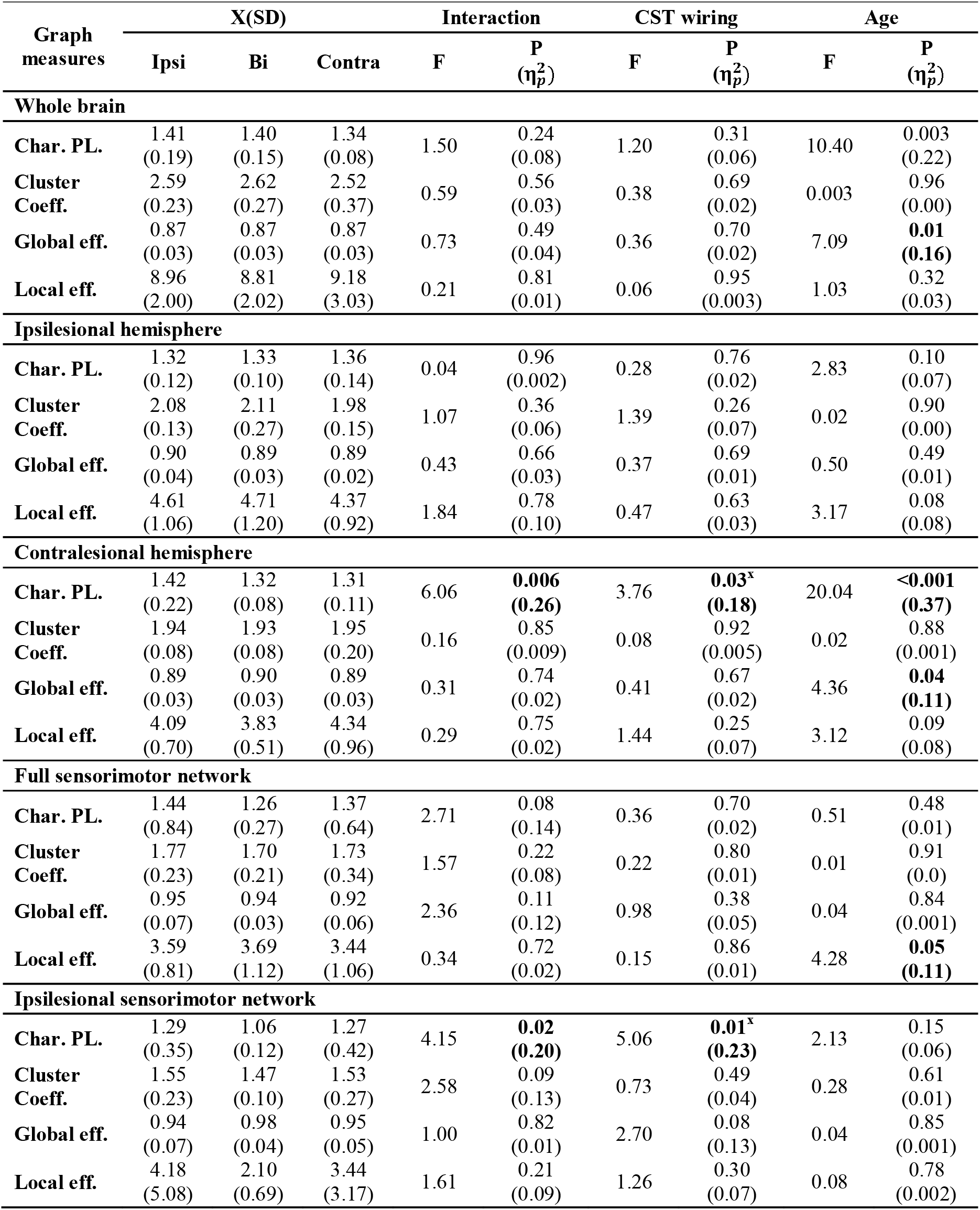
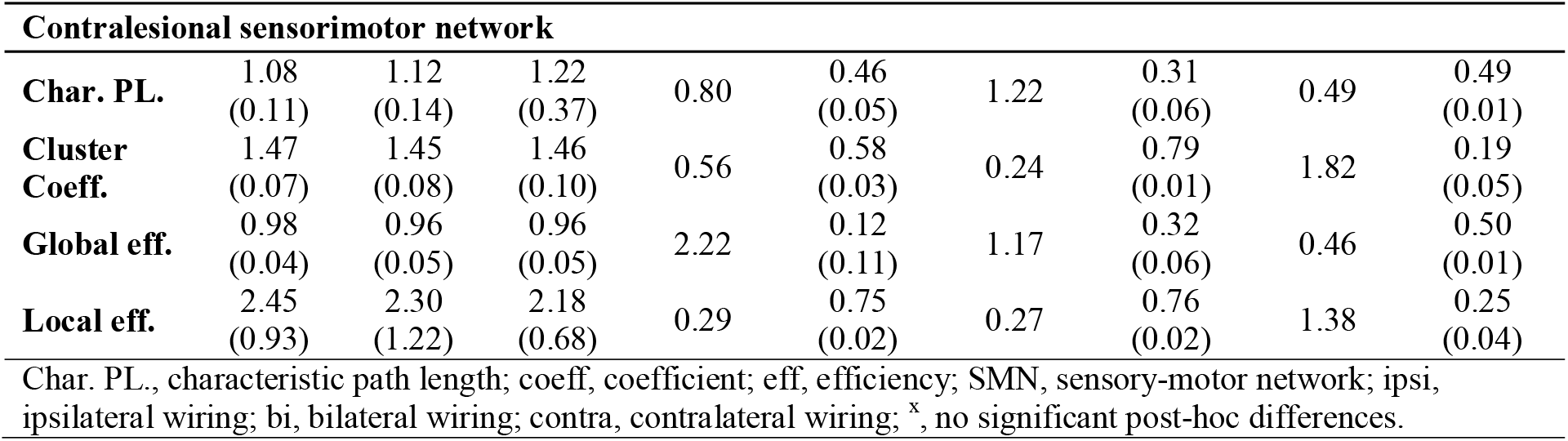
Normalized Graph theory measures across corticospinal tract wiring patterns.

### Elastic-net regularized regression

For this analysis, 39 children with the complete data set were included. One child had missing data for two-point discrimination and stereognosis and 6 children did not undergo the TMS assessment.

The elastic-net regularized regression selected a full LASSO-regression (⍰=1) for four dependent variables (i.e. AHA, grip force of both hands and JTHFT of the dominant hand), while for both somatosensory outcomes a full RIDGE-regression was selected (⍰=0). For the JTHFT of the impaired hand a combination of LASSO and RIDGE regression was used (⍰=0.17). R^2^ was substantial for all models, except for movement duration (i.e. JTHFT). A detailed overview of the output can be found in Table D in Supplementary Materials.

For *bimanual performance*, i.e. AHA, the R^2^ was 0.73 (RMSE=0.56). The CST-wiring pattern was the strongest predictor with a moderate (d=-0.78) to large (d=-0.98) effect size. Compared to children with a contralateral CST-wiring pattern, having an ipsilateral CST-wiring pattern was related to a 0.98xSD lower score on the AHA on average and having a bilateral CST-wiring pattern with a 0.78xSD lower score on average. The other retained variables contributed with a tiny (|d|<0.1) to very small (|d|=0.1-0.2) effect.

The R^2^ for *grip force of the impaired hand* was 0.74 (RMSE=0.54). The CST-wiring pattern was the strongest predictor with moderate negative effect sizes (d=-0.71 and -0.54), with lower values for children with ipsilateral or bilateral CST-wiring patterns compared to those with contralateral CST-wiring patterns. Additionally, age had a small positive effect (d=0.38) on grip force, and local efficiency of the full SMN further contributed with a small negative effect to the model (d=-0.24). Other retained variables only had a very small to tiny effect (|d|<0.12). Age was the only predictor to explain the variance in *grip force of the dominant hand* with a moderate effect (d=0.54).

For *movement duration* (i.e. Jebsen-Taylor Hand function test, JTHFT), the R^2^ was 0.39 (RMSE=0.89) for *the impaired hand* and 0.40 (RMSE=0.83) for the *dominant hand*. Age had a small negative effect (d=-0.22) for movement duration of the dominant hand. All other retained variables contributed with a tiny effect (|d|<0.13).

For both *somatosensory outcomes* all variables were included in the model (⍰=0), suggesting that all variables equally contributed to variation of the dependent variable. However, overall individual effect sizes were tiny to very small (d<0.20). For two-point discrimination the R^2^ was 0.80 (RMSE=0.51), and for stereognosis, the R^2^ was 0.87 (RMSE=0.41)

## Discussion

In this exploratory study, we used graph theory to explore structural brain connectomes across the whole brain in children with uCP, and investigated the relation with upper limb sensory-motor function. We found that structural connectomes were mainly lesion type dependent, whilst they appeared to be similar across different CST-wiring patterns. However, the CST-wiring pattern remained the main predictor for motor function. For somatosensation, there was an equal contribution of all predictors.

A previous study showed that in children with uCP, CDGM lesions damages more brain regions and are more extended compared to PWM lesions.^41^ Also white matter tracts, such as the CST and medial lemniscus, appear to be more severely damaged in children with CDGM lesions compared to children with PWM lesions.^42^ Our findings additionally suggests that children with CDGM lesions have a hyperconnectivity pattern between neighbouring nodes (i.e. increased clustering coefficient) in the ipsilesional hemisphere and SMN which coincided with a decreased capacity in the full SMN connectome to communicate between remote nodes (i.e. increased characteristic path length) compared to children with PWM lesions. CDGM lesions predominantly affect the target sites of the white matter connections requiring those connections to take a detour to communicate between nodes, explaining the increased characteristic path length. Subsequently, as more direct connections between remote nodes are damaged, neighbouring nodes might tend to remain connected to accomodate alternative pathways. This hyperconnectivity pattern also fits within the neural group selection theory that indicates that children with brain damage experience difficulties with selecting the most appropriate neurons to perform a motor task, and will rather select different groups of neurons every trial they perform the same motor task.^43^ This results in the retention of abundant connections rather than pruning towards the most optimal solution. Hence, based on our findings, we hypothesise that maturational processes of pruning and myelination occur more aberrantly in children with CDGM lesions compared to PWM lesions which appears to persist into childhood and adolescence.

This hypothesis is strengthened by the interaction effects found between lesion type and age for some graph metrics. With age, global efficiency in the whole brain and in the contralesional hemisphere and SMN decreased more in children with CDGM lesions compared to PWM lesions, while characteristic path length in the whole brain increased more in children with CDGM lesions. In neurotypical development, global efficiency increases shortly after birth while both clustering coefficient and characteristic path length decreases, a process that continues even into adulthood.^44^ Hence, we hypothesize that the structural connectome of children with CDGM lesions deteriorates more with age compared to children with PWM lesions. Longitudinal studies will be needed to confirm this hypothesis.

So far, only one other research group has used GT-analysis to investigate structural connectomes in children with uCP following perinatal stroke, but only addressed the contralesional hemisphere.^45,46^ Similar to our findings, they found a higher global and local efficiency of the contralesional hemisphere and higher clustering coefficient of the contralesional SMN in children with arterial ischemic stroke (mainly affecting cortical and deep grey matter structures) compared to children with periventricular venous infarction (mainly affecting the white matter) as well as in both patient groups compared to neurotypical developing peers.^45,46^ Nevertheless, since up to fifty percent of children with uCP have bilateral brains lesions,^41^ future research is required to investigate to what extent these changes in the contralesional hemisphere are the result of such bilateral brain damage or rather reflect potential compensatory mechanisms.

Strikingly, only limited differences in structural connectomes were found between CST-wiring patterns. This was an unexpected finding, since reorganisation of the CST is the most well-known example of brain plasticity in children with uCP. CST-projections descend from the brain to the spinal cord both contralateral as well as ipsilateral.^47^ In neurotypical development the ipsilateral projections are gradually withdrawn, while the presence of a (unilateral) lesion may cause the contralateral CST-projections to withdraw and the ipsilateral projections to preserve.^47^ Nevertheless, our findings suggest that the type of CST-wiring pattern occurs independently of how the rest of the brain is structurally reorganised in children with uCP, and that the structural connectome is more determined by the type of the lesion.

In agreement with a body of literature, our elastic-net regularised regression revealed that the type of CST-wiring pattern remains the main determinant of motor function, in particular for bimanual performance and grip force of the impaired hand. Previous studies, including multiple brain lesion characteristics, have also shown the dominance of the CST-wiring pattern in predicting upper limb motor function.^35,42^ For grip force of the impaired hand, age and local efficiency of the full SMN additionally contributed with a small effect size. More specifically, our results suggest that grip force is lower in children with higher local efficiency of the full SMN. This is in line with Craig et al.^45^ who also reported that a higher local efficiency of the contralesional hemisphere was associated with lower motor function. An increase in age further resulted in an increase in grip force.^48^ Furthermore, only age predicted grip force of the dominant hand with a moderate effect size. For movement duration, the lowest R^2^ was found with individual effect sizes that were mostly tiny. For somatosensation all variables contributed to the model. However, although a substantial prediction was achieved, the individual effect sizes of each variable were tiny to very small, indicating that our model could not identify a single predictor that best explains somatosensation.

This study also warrants some critical reflections. First, we this dataset did not include a typical developing group. However, our results, at least for the contralesional side, are in line with the studies by Craig et al.^45,46^ who additionally included a control group. Secondly, the heterogeneity of the included lesions and effects of motion in this paediatric cohort is challenging for conducting diffusion MRI analyses. We coped with these issues by developing a tailored processing pipeline, minimising potential biases. Thirdly, the elastic-net regularized regression was performed on the whole data set, increasing the risk of overfitting the models. Nevertheless, the aim of this research question was to exploratively identify predictors. Finally, we did not correct for multiple testing due to the explorative nature of our study. However, effect sizes were additionally reported and strengthened the statistical findings. Moreover, the results and hypotheses made in this exploratory study could support future studies.

To the best of our knowledge this is the first study that explored the structural connectome using GT-analysis of both the contralesional and ipsilesional hemisphere in children with uCP using a semi-automated analysis. A major strength of this study is that our diffusion MRI sequence allowed for multi-shell multi-tissue CSD-analysis which accomodates the modelling of crossing fibre populations and contributions from different brain tissues.^29^ Moreover, when studying the brain in children with CP using tractography, studies often lose data due to extensive lesions which then results in an underrepresentation of those children. Here, we did not lose any participant data due to the inability of tracking specific tracts owed to the use of VBG^10^ and the graph theoretical framework accounting for the effects of lesions.

In conclusion, our study demonstrated the feasibility of an automated GT-analysis in children with uCP, including children with large lesions. Our results suggest a hyperconnectivity pattern between neighbouring nodes in the ipsilesional hemisphere and SMN. Additionally, the structural connectome did not differ between CST-wiring patterns. However, the CST-wiring pattern outweighed structural connectomes in predicting upper limb motor function, underlining the importance to include this variable when studying structure-function relation for the upper limb in children with uCP. For somatosensation, we could not identify a strong individual predictor. Nevertheless, graph theory analysis seems to be a powerful research tool to strengthen our insights regarding the impact of brain damage on both structural and functional connectomes of the developing brain in children with uCP and how this relates to function, as well as to capture brain changes after intensive therapy models.

## Acknowledgements

The authors would like to thank Dr. Ron Peeters and Jasmine Hoskens for their contributions to this study. We would also like to acknowledge, Prof. Dr. Thomas Neyens and Prof. Dr. Ariel Alonso Abad from the Leuven Biostatistics and Statistical Bioinformatics Centre (L-BioStat), KU Leuven for their advice regarding the statistical analyses. We especially thank all families participating in this study.

