## supplementary methods for "Exploring structural connectomes in children with unilateral cerebral palsy using graph theory"

### Image Processing

Intermodality registration was done using ANTs based on the antsRegistration command used in antsIntermodalityIntrasubject.sh registration script as follows:

antsRegistration -d 3 -o path/2/T1_2_firstshvol_ -m MI[ path/2/wmodf_firstsh.nii.gz , path/2/T1_brain_in.nii.gz ,1,32,Regular,0.25] -c [1000x500x250x0,1e-7,5] -t Affine[0.1] -f 8x4x2x1 -s 4x2x1x0 -u 1 -m mattes[ path/2/wmodf_firstsh.nii.gz, path/2/T1_brain_in.nii.gz,1,32] -c [50x50x0,1e-7,5] -t SyN[0.1,3,0] -f 4x2x1 -s 2x1x0mm -u 1 -z 1 --winsorize-image-intensities [0.005,0 0.995] -v

Bias-field correction of the dMRIs was done using ANTs and MRTrix3 with the following syntax:

dwibiascorrect -ants -ants.b [150,3] -ants.c [500x500,0.0] -ants.s 1 -nthreads 9 -force -mask /path/2/dwi_mask.mif /path/2/dwi_preprocessed_cropped.mif /path/2/dwi_fullpreproc.mif

Eddy current correction was done using dwifslpreproc from MRTrix3 which applied FSL’s Eddy’s CUDA implementation with 10 iterations and --repol for slice-wise outlier detection and replacement with the following syntax:

dwipreproc -nthreads 9 -force /path/to/denoised_degibbsed_padded.mif /path/to/dwi_preprocessed.mif -rpe_none -pe_dir AP -eddy_options " --repol --fwhm=10,0,0,0,0,0,0,0,0,0 --slm=linear --niter=10 --cnr_maps --residuals --mporder=20 --slspec=/path/to/slspec.txt " -eddyqc_text /path/to/eddyqc -nocleanup -tempdir /path/to/eddyqc

The tissue probability maps used for Anatomically-Constrained tractography^1^ were generated using ANTs Atropos-based tissue segmentation^2^, while excluding the lesion from the brain mask. Resulting tissue maps were cleaned to eliminate any falsely labelled voxels by masking the grey matter maps with a twice dilated binary mask of the subject-specific parcellation maps generated by recon-all, then applying a minimum threshold of 0.1. The clean tissue maps were then combined into a 4D map and the lesion mask was appended as the last volume, for the pathological tissue type.

Atropos -d 3 -c [5,0.001] -a path/to/T1_brain_in.nii.gz -i PriorProbabilityImages[4, path/to/wprior_in_pat%d.nii.gz,0.1] -k Gaussian -m [0.3,1,1,1] -o [path/to/atropos_tpms_segmentation.nii.gz,path/to/atropos_tpms_%d.nii.gz] -p Socrates[1] -x path/to/brain_mask_woLesion.nii.gz -v 1

The VBG recon-all generated Desikan-Killiany parcellation maps (with lesion excluded) were propagated through the binary mask of the clean grey matter tissue probability maps. UKBB-derived brainstem VOIs were added to the propagated grey matter parcellation maps, and resulting modified parcellation maps were used to create the subject-specific structural connectomes.

ImageMath 3 path/to/aparc_LC_prop2GM.nii.gz PropagateLabelsThroughMask path/to/GM_mask_clean.nii.gz path/to/VBG_output/aparc_minL_LC.nii.gz
