## Supplementary Material for "Exploring structural connectomes in children with unilateral cerebral palsy using graph theory"

| **Table A. Overview of the nodes used in the SMN subnetwork definition.** | | | |
| --- | --- | --- | --- |
| Region | Fulll SMN | Left SMN | Right SMN |
| Cortex left paracentral | X | X |  |
| Cortex left postcentral | X | X |  |
| Cortex left precentral | X | X |  |
| Cortex left rostral middle frontal | X | X |  |
| Cortex left superior frontal | X | X |  |
| Cortext left superior parietal | X | X |  |
| Cortex left supramarginal | X | X |  |
| Left cerebellum cortex | X |  | X |
| Left thalamus proper | X | X |  |
| Cortex right paracentral | X |  | X |
| Cortex right postcentral | X |  | X |
| Cortex right precentral | X |  | X |
| Cortex right rostral middle frontal | X |  | X |
| Cortex right superior frontal | X |  | X |
| Cortex right superior parietal | X |  | X |
| Cortex right supramarginal | X |  | X |
| Right cerebellum cortex | X | X |  |
| Right thalamus proper | X |  | X |
| Left CST (from UK BioBank) | x |  | X |
| Left ML (from UK BioBank) | X |  | X |
| Right CST (from UK BioBank) | X | X |  |
| Right ML (from UK BioBank) | X | X |  |

| **Table B.1 Participant’s characteristics across lesion types.** | | |
| --- | --- | --- |
|  | PWM lesions (N=28) | CDGM lesions (N=18) |
| **Sex** |  |  |
| Male | 15 | 12 |
| Female | 13 | 6 |
| **Age** |  |  |
| at MRI | 10y6m (±2y9m) | 10y10m (±2y10m) |
| at clinical assessment | 10y7m (±2y8m) | 10y10m (±2y9m) |
| **Side of hemiplegia** |  |  |
| Right | 14 | 11 |
| Left | 14 | 7 |
| **MACS** |  |  |
| Level I | 12 | 3 |
| Level II | 12 | 4 |
| Level III | 4 | 11 |
| **CST wiring pattern** |  |  |
| contralateral | 10 | 2 |
| bilateral | 7 | 7 |
| ipsilateral | 8 | 6 |
| PWM, periventricular white matter; CDGM, cortical and deep grey matter; MACS, Manual Ability Classification System; CST, corticospinal tract. | | |

| **Table B.2 Participant’s characteristics across lesion types.** | | | |
| --- | --- | --- | --- |
|  | PWM lesions (N=28) | CDGM lesions (N=18) | P-value |
| **Grip force DH**^a^ (kilograms) | 17.92 (±7.39) | 17.06 (±6.27) | 0.358 |
| **Grip force nDH**^a^ (kilograms) | **9.55 (±5.92)** | **5.66 (±3.67)** | **0.003*** |
| **2-point discrimination**^b^  (0-11mm) | **3 [2-4]** | **11 [7.5-11]** | **<0.001*** |
| **Stereognosis**^b^ (0-6 objects) | **6 [5-6]** | **2 [0.5-5]** | **0.002*** |
| **AHA**^b^ (0-100 AHA-units) | **63.5 [58.25-82.50]** | **55 [48-61.75]** | **0.01*** |
| **JTHFT DH**^b^ (seconds) | 33.830 [29.19-38.74] | 38.31 [30.55-45.11] | 0.131 |
| **JTHFT nDH**^b^ (seconds) | **95.26 [58.37-221.74]** | **310.72 [94.84-452.68]** | **0.03*** |
| PWM, periventricular white matter; CDGM, cortical and deep grey matter; a, ANCOVA (with age as covariate) reported with means ± SD; b, Mann-Whitney U test reported with median [Q1-Q3]; DH, dominant hand; nDH, non-dominant hand; mm, millimetres; AHA, Assisting Hand Assessment; JTHFT, Jebsen-Taylor Hand Function Test. | | | |

| **Table C.1 Participant’s characteristics across CST wiring groups.** | | | |
| --- | --- | --- | --- |
|  | Contralateral (N=12) | Bilateral (N=14) | Ipsilateral (N=14) |
| **Sex** |  |  |  |
| Male | 7 | 7 | 8 |
| Female | 5 | 7 | 6 |
| **Age** |  |  |  |
| at MRI | 10y7m (±2y11m) | 10y1m (±2y8m) | 10y8m (±2y10m) |
| at clinical assessment | 10y7m (±2y11m) | 10y3m (±2y7m) | 10y9m (±2y9m) |
| **Side of hemiplegia** |  |  |  |
| Right | 7 | 5 | 9 |
| Left | 5 | 9 | 5 |
| **MACS** |  |  |  |
| Level I | 10 | 2 | 1 |
| Level II | 2 | 8 | 6 |
| Level III | 0 | 4 | 7 |
| **Lesion type** |  |  |  |
| PWM lesions | 10 | 7 | 8 |
| CDGM lesions | 2 | 7 | 6 |
| PWM, periventricular white matter; CDGM, cortical and deep grey matter; MACS, Manual Ability Classification System; CST, corticospinal tract. | | | |

| **Table C.2 Participant’s characteristics across CST wiring groups.** | | | | |
| --- | --- | --- | --- | --- |
|  | Contralateral (N=12) | Bilateral (N=14) | Ipsilateral (N=14) | P-value* |
| **Grip force DH**  (kilograms) | 14.33 [12.62-22.75] | 16.33 [9.25-21.5] | 18.67 [14-22.03] | 0.771 |
| **Grip force nDH**  (kilograms) | 3.57 [2.25-8.75] | 4.83 [3.58-7] | 14.08 [8.33-16.42] | **<0.001*^,a,b^** |
| **2-point discrimination**  (0-11mm) | 5 [3-11] | 3.5 [2-11] | 3 [2-3.75] | 0.097 |
| **Stereognosis**  (0-6 objects) | 4 [2.5-6] | 4.5 [0.75-6] | 6 [6-6] | **0.001*^,a,b^** |
| **AHA**  (0-100 AHA-units) | 55 [48.75-66.25] | 57.5 [54.5-60.75] | 84 [80.25-87] | **<0.001*^,a,b^** |
| **JTHFT DH** (seconds) | 37.26 [28.75-42.02] | 33.42 [29-38.33] | 34.64 [28.55-45.93] | 0.800 |
| **JTHFT nDH** (seconds) | 232.78 [87.08-397.78] | 168.33 [102.51-253.65] | 63 [41.24-81.12] | **0.001*^,a,b^** |
| *, Kruskall-Wallis test reported with median [Q1-Q3]; a, post-hoc significance contralateral versus ipsilateral; b, post-hoc significance bilateral versus contralateral; DH, dominant hand; nDH, non-dominant hand; mm, millimetres; AHA, Assisting Hand Assessment; JTHFT, Jebsen-Taylor Hand Function Test. | | | | |

| **Table D. Results of the elastic net regularised regression** | | | | | | | | |
| --- | --- | --- | --- | --- | --- | --- | --- | --- |
| **Predictors** | | **Grip force impaired hand** | **Grip force dominant hand** | **2PD** | **STEREO** | **JTHFT impaired hand** | **JTHFT dominant hand** | **AHA** |
| **Lamda value** | | 0.06 | 0.23 | 0.95 | 0.62 | 1.32 | 0.24 | 0.06 |
| **Alpha value** | | 1 | 1 | 0 | 0 | 0.17 | 1 | 1 |
| **Classical neurological predictors** | **age** | 0.38 | 0.54 | -0.01 | 0.11 |  | -0.22 | 0.0009 |
|  | **Lesion volume** | -0.02 |  | 0.06 | -0.06 | 0.05 |  | -0.11 |
|  | **Lesion type** |  |  | -0.14 | 0.12 |  |  |  |
|  | **CST_1** | -0.71 |  | 0.02 | -0.03 |  |  | -0.98 |
|  | **CST_2** | -0.54 |  | 0.04 | -0.07 |  |  | -0.78 |
| **Whole brain** | **Norm. Char. PL.** |  |  | 0.08 | -0.1 |  |  |  |
|  | **Norm.Cluster coeff.** | -0.06 |  | 0.06 | -0.07 |  |  |  |
|  | **Norm.Global eff.** |  |  | 0.02 | -0.07 |  |  |  |
|  | **Norm.Local eff.** |  |  | 0.06 | -0.06 |  |  |  |
| **Ipsilesional hemisphere** | **Norm.Char. PL.** |  |  | -0.09 | 0.13 | -0.06 | -0.01 | 0.23 |
|  | **Norm.Cluster coeff.** |  |  | 0.12 | -0.20 | 0.05 | 0.004 | -0.12 |
|  | **Norm.Global eff.** |  |  | 0.03 | -0.07 |  |  |  |
|  | **Norm.Local eff.** |  |  | 0.12 | -0.19 |  |  |  |
| **contralesional hemisphere** | **Norm.Char. PL.** |  |  | 0.05 | -0.08 |  |  |  |
|  | **Norm.Cluster coeff.** |  |  | -0.02 | 0.08 |  |  |  |
|  | **Norm.Global eff.** |  |  | 0.02 | -0.09 | 0.03 | 0.13 |  |
|  | **Norm.Local eff.** | 0.12 |  | 0.04 | 0.04 |  |  |  |
| **Full SMN** | **Norm.Char. PL.** |  |  | 0.07 | -0.0009 | 0.01 |  |  |
|  | **Norm.Cluster coeff.** |  |  | 0.03 | -0.04 |  |  |  |
|  | **Norm.Global eff.** | -0.004 |  | 0.12 | -0.11 |  |  |  |
|  | **Norm.Local eff.** | -0.24 |  | 0.07 | -0.13 | 0.06 |  | -0.13 |
| **Ipsilesional SMN** | **Norm.Char. PL.** |  |  | 0.06 | 0.04 |  |  |  |
|  | **Norm.Cluster coeff.** |  |  | 0.08 | -0.04 |  |  |  |
|  | **Norm.Global eff.** |  |  | 0.06 | -0.07 |  | -0.05 |  |
|  | **Norm.Local eff.** |  |  | 0.01 | 0.02 |  | -0.05 |  |
| **contralesional SMN** | **Norm.Char. PL.** | 0.01 |  | -0.11 | 0.08 |  |  | 0.04 |
|  | **Norm.Cluster coeff.** | 0.11 |  | -0.05 | 0.08 |  |  |  |
|  | **Norm.Global eff.** |  |  | 0.1 | -0.09 |  |  |  |
|  | **Norm.Local eff.** |  |  | 0.15 | -0.09 |  |  |  |
| **Outcome model** | **R²** | 0.70 | 0.54 | 0.73 | 0.82 | 0.19 | 0.29 | 0.68 |
|  | **RMSE** | 0.54 | 0.67 | 0.51 | 0.41 | 0.89 | 0.83 | 0.56 |
| 2PD, two-point discrimination; STEREO, stereognosis; JTHFT, Jebsen-Taylor Hand Function test; AHA, Assisting Hand Assessment; Norm., Normalized; Char. PL., characteristic path length; coeff, coefficient; eff, efficiency; SMN, sensory-motor network; CST_1, ipsilateral compared to contralateral; CST_2, bilateral compared to contralateral; RMSE, root mean square error; Green, large effect size; Yellow, medium effect size; blue, small effect size. | | | | | | | | |
