## Supplementary Figures for "Exploring structural connectomes in children with unilateral cerebral palsy using graph theory"

***Figures S1-S5. Lesion types.***

**Figure S1. Scatter plot of the interaction between age and lesion type for normalized global efficiency of the whole brain.**

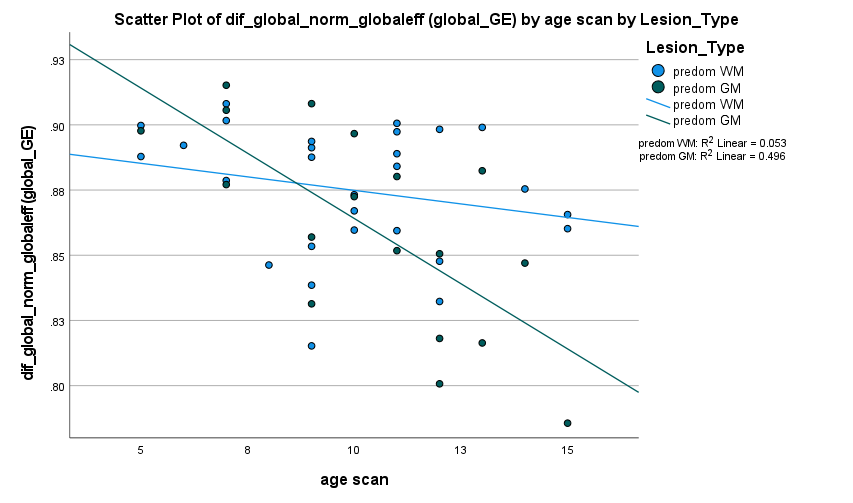

Legend: x-axis, age at time of the MRI-scan; y-axis, normalized global efficiency whole brain.

**Figure S2.** **Scatter plot of the interaction between age and lesion type for normalized global efficiency of the contralesional hemisphere.**

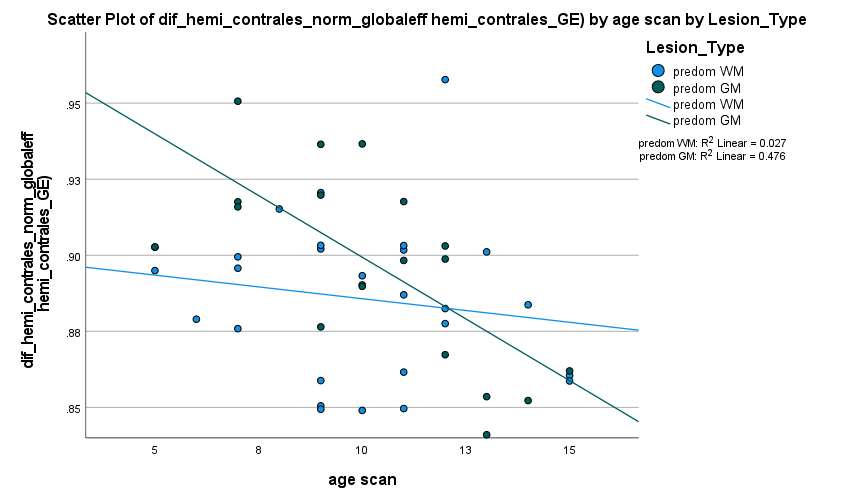

Legend: x-axis, age at time of the MRI-scan; y-axis, normalized global efficiency contralesional hemisphere.

**Figure S3. Scatter plot of the interaction between age and lesion type for normalized global efficiency of the contralesional SMN.**

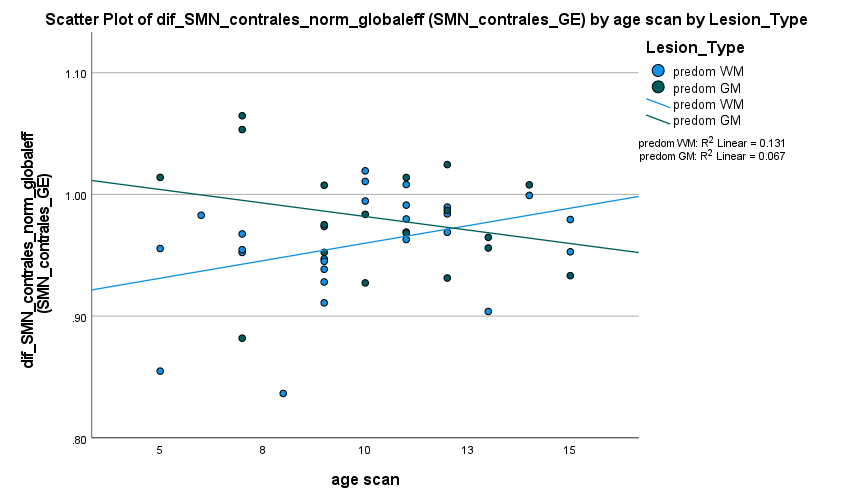

Legend: x-axis, age at time of the MRI-scan; y-axis, normalized global efficiency contralesional sensorimotor network.

**Figure S4. Scatter plot of the interaction between age and lesion type for normalized characteristic path length of the whole brain.**

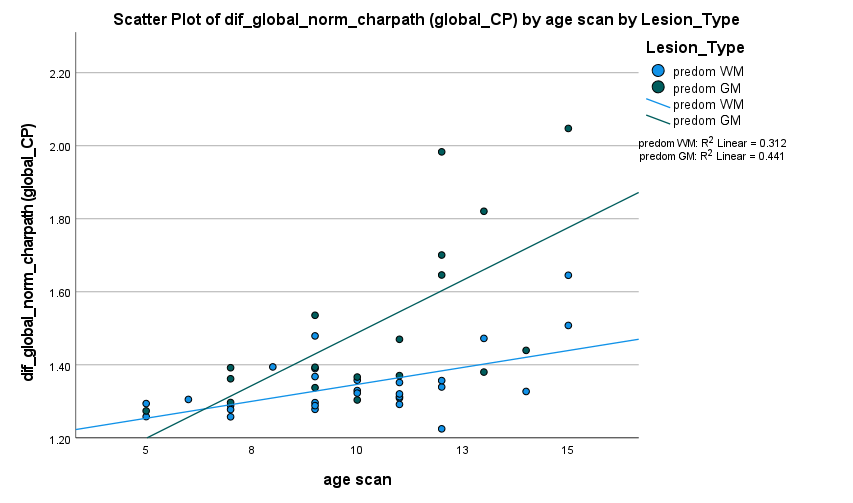

Legend: x-axis, age at time of the MRI-scan; y-axis, normalized characteristic path length whole brain.

**Figure S5. Scatter plot of the interaction between age and lesion type for normalized characteristic path length of the ipsilesional hemisphere.**

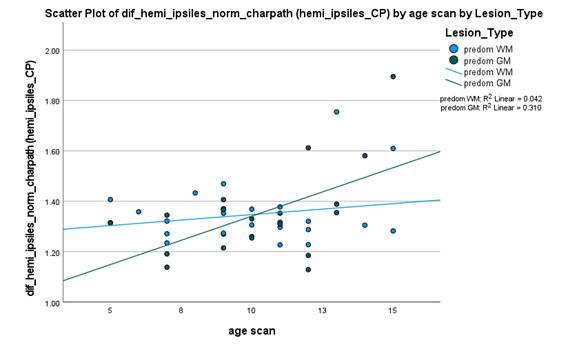

Legend: x-axis, age at time of the MRI-scan; y-axis, normalized characteristic path length ipsilesional hemisphere.

***Figures S6-S7. CST wiring groups.***

**Figure S6. Scatter plot of the interaction between age and CST wiring patterns for normalized characteristic path length of the contralesional hemisphere.**

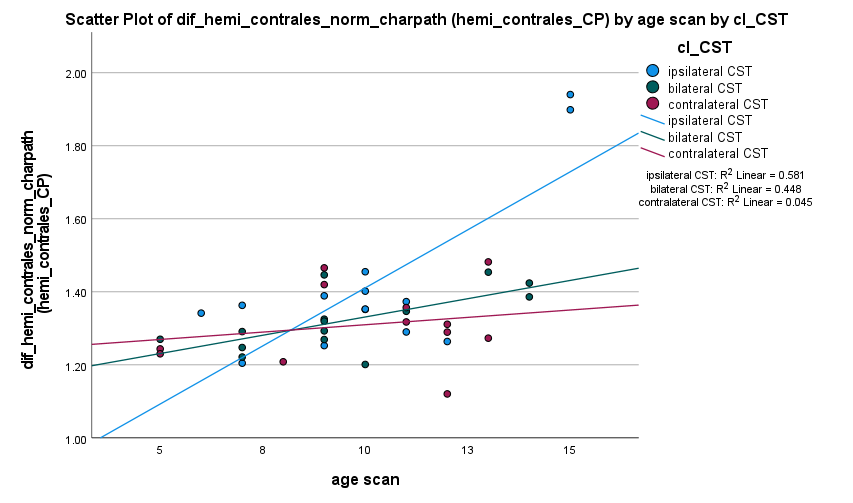

Legend: x-axis, age at time of the MRI-scan; y-axis, normalized characteristic path length contralesional hemisphere; cl_CST, CST wiring groups.

**Figure S7. Scatter plot of the interaction between age and CST wiring patterns for normalized characteristic path length of the ipsilesional SMN.**
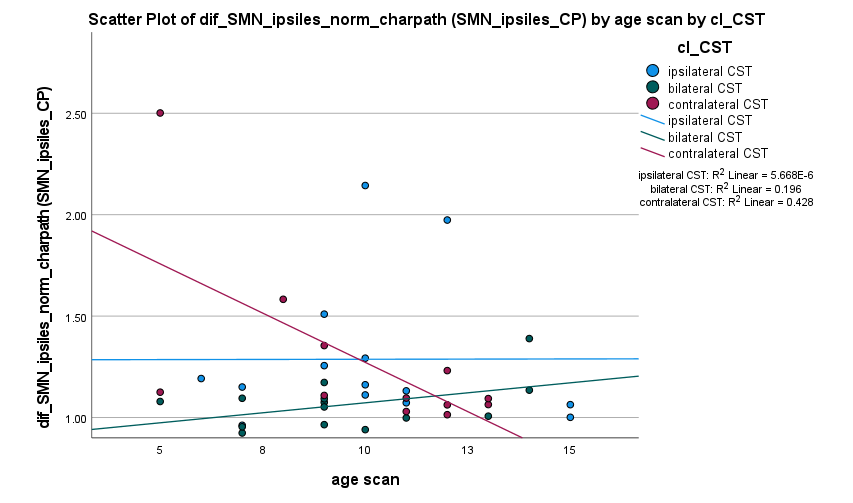

Legend: x-axis, age at time of the MRI-scan; y-axis, normalized characteristic path length ipsilesional sensorimotor network; cl_CST, CST wiring groups.
